# Comparing Machine and Deep Learning Models for Pediatric Anxiety Classification using Structured EHRs and Area-based Measures of Health Data

**DOI:** 10.1101/2025.05.01.25326789

**Authors:** Eric W. Lee, Sanghyun Choo, Dakotah Maguire, Abhishek Shivanna, Daniel Santel, Surbhi Bhatnagar, Ian Goethert, Kelly Patterson, Jay Gholap, Heidi A. Hanson, Mayanka Chandrashekar, Robert T. Ammerman, John P. Pestian, Tracy Glauser, Cole Brokamp, Jeffrey R. Strawn, Anuj Kapadia, Greeshma Agasthya

## Abstract

**Objective:** This study investigates the performance of various machine learning (ML) and deep learning (DL) models to classify pediatric patients at risk of anxiety disorders using electronic health records (EHRs). By leveraging EHR data and including Area-based measures of health (ABMH) data, this approach aims to enable proactive care by monitoring potential anxiety onset comprehensively across various age groups.

**Methods:** In this study, we trained a series of ML and DL models to classify youth at risk of developing anxiety disorders. ML models (Logistic Regression, Decision Tree, Random Forest, K-Nearest Neighbors, XGBoost) and DL models (LSTM, GRU, RETAIN, Dipole) were trained using structured EHR data from 30-day periods before anxiety diagnoses. Two datasets per age group were used: one with structured EHR data only and another with incorporating both structured EHR and ABMH data. Model performance was assessed using accuracy, the AUROC, AUPRC, PPV, NPV, and F1 scores.

**Results:** The ML models provided a solid performance baseline, with XGBoost showing strong baseline performance across age groups, with AUROC scores of 0.817 (structured EHR) and 0.816 (structured EHR + ABMH). Between DL models, RETAIN and Dipole performed the best. For example, RETAIN achieved AUROC scores of 0.851 (structured EHR) and 0.853 (structured + ABMH), while Dipole scored 0.853 and 0.857, respectively, for 8-year-olds. These results underscore the viability of both ML and DL models for the early detection of pediatric anxiety disorders.

**Conclusion:** This study comprehensively investigated ML and DL models for diagnosing pediatric anxiety. We demonstrated that ML and DL models can effectively monitor probable anxiety onset within an EHR system and also with the ABMH data. We discovered that model performance varied with age, indicating the need for personalized model development per age group for effective clinical predictive analytics.

## Introduction

Anxiety disorders are the most common type of mental disorder, and an estimated 19.1% of adults in the U.S. population have an anxiety disorder [1]. Anxiety disorders typically begin during childhood or adolescence and persist into adulthood, with a lifetime prevalence [2]. They manifest at an earlier stage of development compared to depression [3] and, if not treated, are associated with significant short- and long-term impairment [4]. Early-onset anxiety disorders are more likely to lead to significant depression, substance dependency, suicidal behavior, and educational underachievement [5]. Therefore, implementing effective strategies for the early identification of anxiety could result in improved health outcomes throughout an individual’s lifespan.

Currently, the identification of children at risk for developing anxiety disorders is sub-optimal. It is estimated that 80% of youth with diagnosable anxiety are not getting treatment [6]. Traditionally, anxiety is diagnosed by pediatricians, primary care clinicians, and psychiatric clinicians and requires knowledge of the patient’s history, specific symptoms (e.g., sleep patterns, concentration, restlessness, etc), and physical health. Given the extensive nature of medical records, it is often difficult for a clinician to ingest and summarize the large volume of information about a patient’s health across their life course [7]. This is a significant barrier to effective and timely intervention, impacting short- and long-term patient outcomes. Enhancing electronic health records (EHRs) systems with intelligent, streamlined tools for early diagnoses of clinical anxiety using classification models, i.e., machine learning (ML) and deep learning (DL), could substantially improve clinical decision-making processes and reduce the likelihood of missed diagnoses. Furthermore, early and effective anxiety diagnosis can improve the long-term health of the patient and diminish long-term healthcare expenses associated with a missed diagnosis [8]. In this study, we compare the performance of several ML and DL methods in identifying and classifying pediatric patients with anxiety. These identification and classification tasks are performed using time-dependent and static features in the EHR, along with Area-based measures of health (ABMH) data for ages 2 to 21.

EHRs contain structured data such as diagnosis and treatment codes, prescription medications, and demographic data, which researchers have utilized to build ML and DL models to predict illness and diseases [9–11]. Although successful, many classification models ignore the time-dependent EHR data, which can be used effectively as a signal of future risk [12]. Each patient’s diagnoses are documented using the International Classification of Disease (ICD) codes at each clinical encounter, and the number of encounters increases over time. This sparse nature makes using the time-dependent EHR data challenging [13]. Studies have explored different techniques using recurrent neural networks (RNNs)-based models to handle long sequences of encounters [14–17]. Although several of these approaches have been proposed, a comprehensive comparative assessment of the different methods has not yet been performed, and is needed to confirm the best model to identify pediatric anxiety.

The influence of ABMH on child and adolescent development—and the risk of psychopathology—has been well-established over decades of research [3, 18, 19]. Yet, despite this robust evidence, ABMH is inconsistently integrated into our understanding of how and why psychiatric disorders emerge in children and adolescents [20]. Environmental exposures, such as poverty or unsafe neighborhoods as well as neighborhood-level air pollution [21, 22], are linked to a higher likelihood of anxiety and related disorders [19, 23]. Limited family resources significantly increase the risk of mood disorders, while negative life events and caregiver strain not only heighten the risk for anxiety but also reduce the chances of responding to treatment [24, 25]. Additionally, exposure to childhood violence has a clear association with the development of anxiety and depression, with these effects being particularly pronounced in younger children and girls [26]. Yet, despite this, much of our work in understanding and screening for anxiety disorders has focused narrowly on individual risk factors—like inhibited temperament, family history, or subsyndromal symptoms—without adequately considering the influence of social and environmental contexts. This oversight is more than a knowledge gap; it is a missed opportunity. Incorporating ABMH into screening represents a substantial advance in assessing risk and an opportunity to identify vulnerabilities earlier, design more comprehensive interventions, and ultimately reduce disparities in mental health care access and outcomes. For clinicians, screening and treating anxiety disorders in youth means moving closer to a system that recognizes the interconnectedness of their environment, experiences, and biology [27].

This study investigates the utility of various computational methods for identifying pediatric patients with anxiety using time-dependent and static features in EHRs and ABMH data. First, we assess the performance of ML models that incorporate only recent 30-day information with a 30-day blackout period prior to diagnosis. Second, we assess the performance of DL models that incorporate time-dependent features that cover the span of the patient’s history. We evaluate the performance of each model across different age groups with two nested datasets: (1) structured features generated from EHR data (*EHR*) and (2) the combination of structured EHR and ABMH data (*EHR+ABMH*). The overall goal is to provide bioinformaticians and clinicians with comprehensive information that can be used to guide the development of predictive models for pediatric anxiety.

## Methods

This section introduces the datasets used in this study: (1) structured features generated from EHR data and (2) ABMH features extracted from multiple sources and linked to a patient’s residential location at the census tract level. Our approach applies a suite of ML models that are commonly used in classification tasks, including logistic regression (LR), decision tree (DT), random forest (RF) [28], k-nearest neighbors (KNN), and extreme gradient boosting (XGBoost) [29]. For the DL models, we select RNN-based models to handle long sequences of encounters such as gated recurrent unit (GRU) [30], long-short term memory (LSTM) [31], reverse time attention (RETAIN) [16], and diagnosis classification model (Dipole) [17]. In anxiety, the presentation of disease and features important for diagnosis can vary by age, and hence, the models were stratified by age to account for this effect. Fig 1 provides a comprehensive summary of our approach for developing classification algorithms on *EHR* datasets for a single age group.

**Fig 1.**
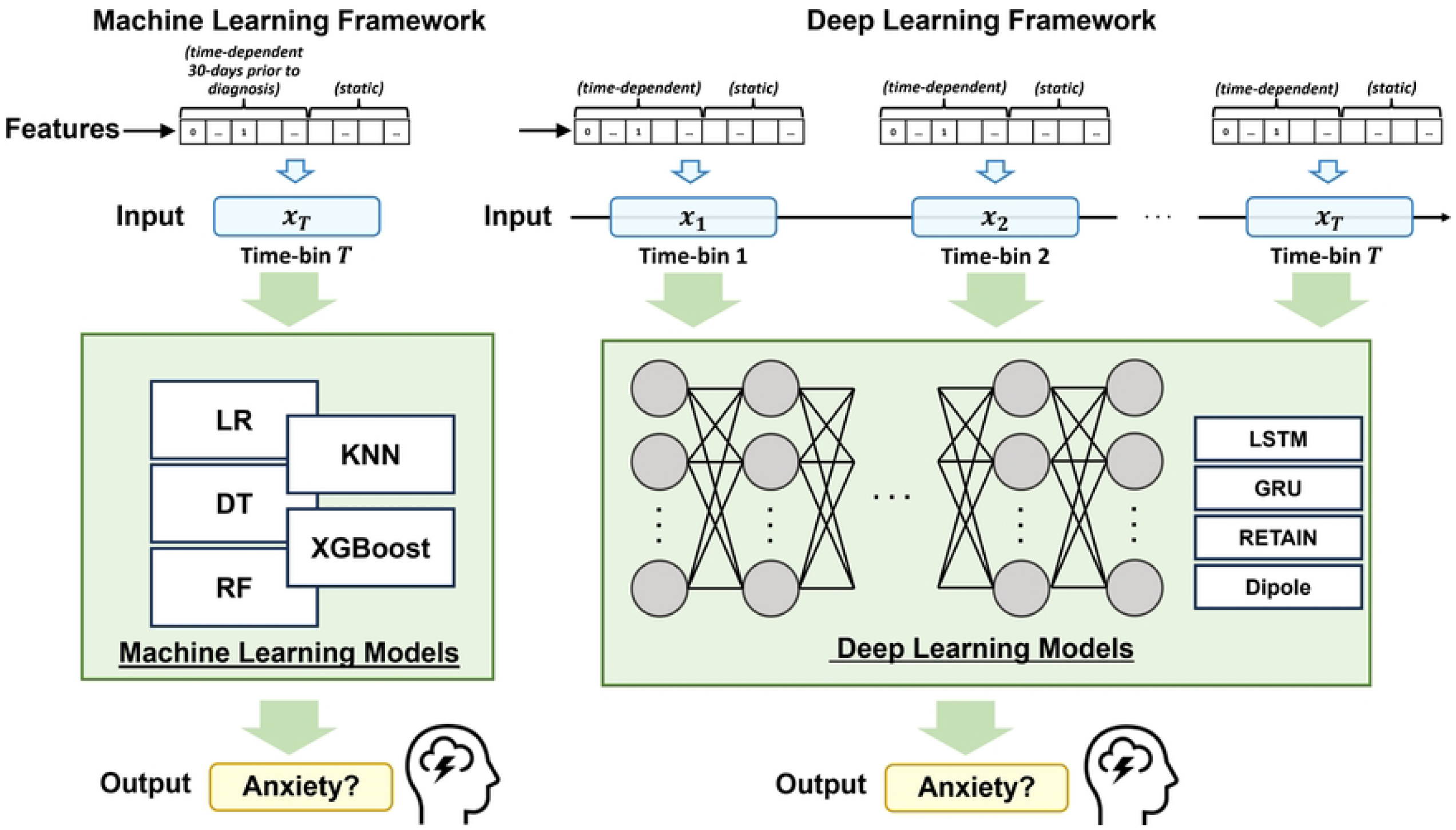
A framework overview for age-stratified pediatric anxiety classification.

### Dataset

The Cincinnati Children’s Hospital Institutional Review Board approved this retrospective study as STUDY(2020-0942). This study uses Cincinnati Children’s Hospital Medical Center’s (CCHMC) pediatric EHRs from 1.3 million patients collected from January 1st, 2009, to March 31st, 2022. The data used in this study was copied by members at CCHMC on April 1, 2022 and processed to create a static foundational database. In May 2022, a copy of the database was driven to Oak Ridge National Laboratory where it was then hosted on a secure platform. The research team accessed the data used in this study from January 2023 to January 2024.

In this study, we used a retrospective case-control study design, where each anxiety case was matched to a control by age at the time of the case’s diagnosis and sex assigned at birth. Anxiety cases were identified using ICD codes described in S1 Table. The date of anxiety onset was determined using the first instance of an anxiety ICD code. This resulted in the selection of 53,728 anxiety patients diagnosed between the ages of 2 and 21 between 2009 to 2022. At least 1 visit in the 18 months prior to the diagnosis date was required for a patient to be considered in the case or control selection. For the control selection, the patient was required to be the same sex assigned at birth as the case, born within 30 days of the case, have not developed anxiety at the time of the case’s anxiety diagnosis record, and have had at least one encounter in the EHRs in the 18 months preceding the case’s anxiety diagnosis date. The matched case-control dataset was then stratified by single-year age groups from ages 2 to 21 based on age of diagnosis of the case. The descriptive statistics for each age group are shown in Table 1.

**Table 1.**
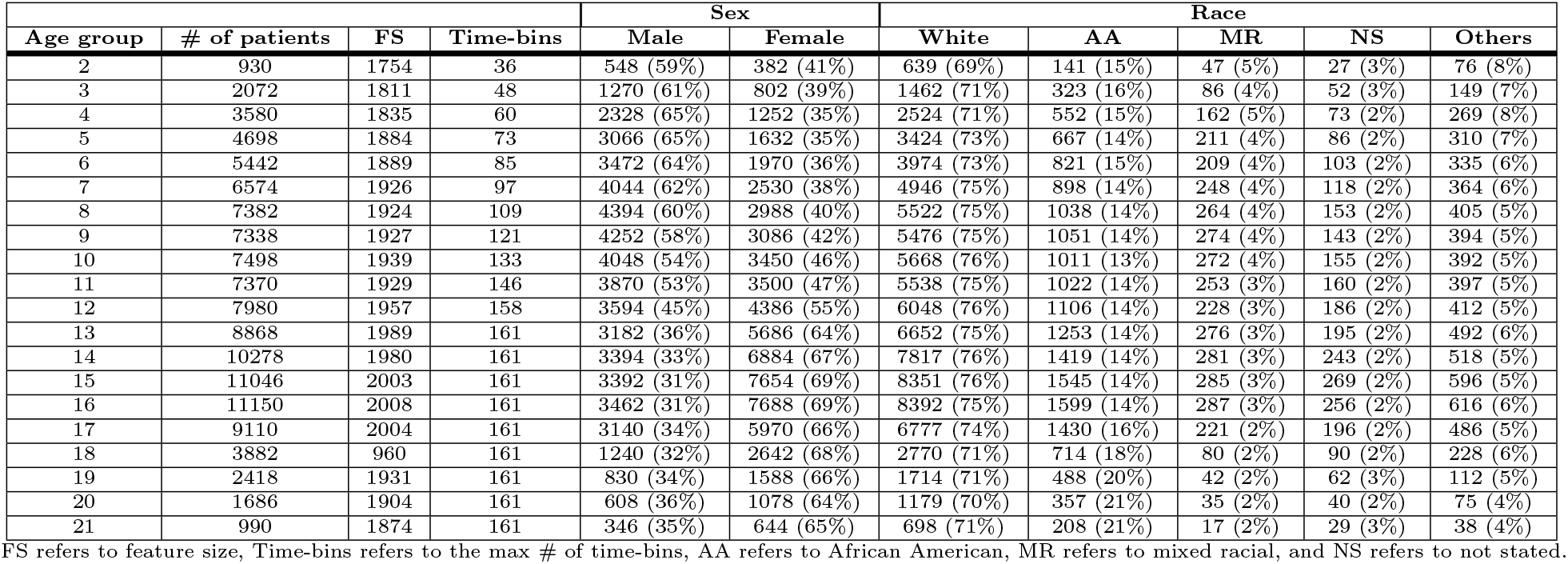
Statistics of the Structured EHR Data.

| Age group | # of patients | FS | Time-bins | Sex |  | Race |  |  |  |  |
| --- | --- | --- | --- | --- | --- | --- | --- | --- | --- | --- |
|  |  |  |  | Male | Female | White | AA | MR | NS | Others |
| 2 | 930 | 1754 | 36 | 548 (59%) | 382 (41%) | 639 (69%) | 141 (15%) | 47 (5%) | 27 (3%) | 76 (8%) |
| 3 | 2072 | 1811 | 48 | 1270 (61%) | 802 (39%) | 1462 (71%) | 323 (16%) | 86 (4%) | 52 (3%) | 149 (7%) |
| 4 | 3580 | 1835 | 60 | 2328 (65%) | 1252 (35%) | 2524 (71%) | 552 (15%) | 162 (5%) | 73 (2%) | 269 (8%) |
| 5 | 4698 | 1884 | 73 | 3066 (65%) | 1632 (35%) | 3424 (73%) | 667 (14%) | 211 (4%) | 86 (2%) | 310 (7%) |
| 6 | 5442 | 1889 | 85 | 3472 (64%) | 1970 (36%) | 3974 (73%) | 821 (15%) | 209 (4%) | 103 (2%) | 335 (6%) |
| 7 | 6574 | 1926 | 97 | 4044 (62%) | 2530 (38%) | 4946 (75%) | 898 (14%) | 248 (4%) | 118 (2%) | 364 (6%) |
| 8 | 7382 | 1924 | 109 | 4394 (60%) | 2988 (40%) | 5522 (75%) | 1038 (14%) | 264 (4%) | 153 (2%) | 405 (5%) |
| 9 | 7338 | 1927 | 121 | 4252 (58%) | 3086 (42%) | 5476 (75%) | 1051 (14%) | 274 (4%) | 143 (2%) | 394 (5%) |
| 10 | 7498 | 1939 | 133 | 4048 (54%) | 3450 (46%) | 5668 (76%) | 1011 (13%) | 272 (4%) | 155 (2%) | 392 (5%) |
| 11 | 7370 | 1929 | 146 | 3870 (53%) | 3500 (47%) | 5538 (75%) | 1022 (14%) | 253 (3%) | 160 (2%) | 397 (5%) |
| 12 | 7980 | 1957 | 158 | 3594 (45%) | 4386 (55%) | 6048 (76%) | 1106 (14%) | 228 (3%) | 186 (2%) | 412 (5%) |
| 13 | 8868 | 1989 | 161 | 3182 (36%) | 5686 (64%) | 6652 (75%) | 1253 (14%) | 276 (3%) | 195 (2%) | 492 (6%) |
| 14 | 10278 | 1980 | 161 | 3394 (33%) | 6884 (67%) | 7817 (76%) | 1419 (14%) | 281 (3%) | 243 (2%) | 518 (5%) |
| 15 | 11046 | 2003 | 161 | 3392 (31%) | 7654 (69%) | 8351 (76%) | 1545 (14%) | 285 (3%) | 269 (2%) | 596 (5%) |
| 16 | 11150 | 2008 | 161 | 3462 (31%) | 7688 (69%) | 8392 (75%) | 1599 (14%) | 287 (3%) | 256 (2%) | 616 (6%) |
| 17 | 9110 | 2004 | 161 | 3140 (34%) | 5970 (66%) | 6777 (74%) | 1430 (16%) | 221 (2%) | 196 (2%) | 486 (5%) |
| 18 | 3882 | 960 | 161 | 1240 (32%) | 2642 (68%) | 2770 (71%) | 714 (18%) | 80 (2%) | 90 (2%) | 228 (6%) |
| 19 | 2418 | 1931 | 161 | 830 (34%) | 1588 (66%) | 1714 (71%) | 488 (20%) | 42 (2%) | 62 (3%) | 112 (5%) |
| 20 | 1686 | 1904 | 161 | 608 (36%) | 1078 (64%) | 1179 (70%) | 357 (21%) | 35 (2%) | 40 (2%) | 75 (4%) |
| 21 | 990 | 1874 | 161 | 346 (35%) | 644 (65%) | 698 (71%) | 208 (21%) | 17 (2%) | 29 (3%) | 38 (4%) |

FS refers to feature size, Time-bins refers to the max # of time-bins, AA refers to African American, MR refers to mixed racial, and NS refers to not stated.

### Data Preprocessing and Feature Engineering

Comprehensive patient histories were extracted from the CCHMC EHR. We then followed a series of preprocessing and feature engineering steps to create the final analytic tables to train the ML and DL models: 1) We extracted the static features that describe patient characteristics that do not change over time. 2) We collapsed time-dependent features into 30-day time-bins to capture the relevant EHR events during each time-bin. 3) We created an analytic file for the ML analyses that only included 30-day information prior to a recent 30-day blackout period to the time of diagnosis. 4) We created a time-dependent dataset for the DL analyses that included 30-day time-bins from birth to the time of the case’s diagnosis with a 30-day blackout period. 5) We appended the ABMH data to the analytic files created in steps three and four. 6) We split the analytic files into age-specific datasets for analysis. 7) We created train/test splits using patient ID.

#### Structured EHR Data

The structured data includes information from two types of features: time-dependent (features that change over time) and static (features that do not change over time). The time-dependent features consist of diagnosis codes, procedure codes, medication codes, visit metadata (encounter type, provider type, place of service, care site, hospitalization), and measures (BMI, height, weight, blood pressure, heart rate). The static features include information set at birth, such as allergies, family mental health history, and patient demographics. The categorization of allergies (food, medications, and environment) and family histories (psychiatric disorders, substance abuse, sexual/verbal abuse, autism, attention-deficit/hyperactivity disorder, and developmental disorder). For the structured EHR data, we use frequency encoding and replace missing values with -1. Detailed information on time-dependent and static features is discussed in S1 Appendix.

#### Generation of Time-dependent Features

Fig 1 illustrates the utilization of time-dependent features. For these experiments, we used 30-day time-bins. The 30-day time-bin consolidates all EHR and ABMH data within each time-bin during the 30-day period. Time-bins *x*_1_, *x*_2_, …, *x*_*T*_ extend from the time of birth to the time of the case’s diagnosis and include relevant EHR events that occurred during each 30-day window. Using the 30-day time-bins results in approximately 12 time-bins per year. We utilize this sequence of time-bins as the input feature for the DL-based models, while for the ML-based models, we use the final time-bin (*x*_*T*_) as the input feature. Table 1 summarizes the feature size and the number of time-bins of each age group.

#### Area-based Measures of Health (ABMH) Data

Measuring ABMH across time using EHR data is a challenging task. This information is not consistently stored in EHR records and must be reconstructed using residential history information for each patient. In order to do this effectively, each residential address must be geocoded, assigned a relevant time window, and spatially joined to external sources of data that describe a patient’s community environment. We use the DeGAUSS package [32] to define a community environment at a single time point and then develop ABMH trajectories using a patient’s residential history.

We sought to capture the dynamic ABMH a patient might be exposed to throughout their life course. Therefore, ABMH features were considered to be time-dependent. This was done by constructing patient residential histories from residential address information captured in the CCHMC enterprise data warehouse (EDW). We extracted all known residential locations for each patient and assigned a start and stop date for each unique location. The date of visit corresponding to the residential location was used to construct the residential history of the patient from birth to the time of anxiety diagnosis. This required several assumptions: (1) the patient’s first observed address in the EHR record was also their address at the time of birth, and (2) the patient resided at their last known or current location for the full length of time between visits. DeGauss was used to geocode and link each patient’s residential location to a corresponding United States Census Tract (CT) at a single point in time. We accounted for changes in US Census Tract boundaries over time to ensure a correct characterization of the community a patient resided in at each point in time. The CT level measures used to characterize a patient’s ABMH over time can be found in S2 Table. Using a modified version of the time-bin code, we created environment measures for every 30-day time-bin used in the EHR data construction. If an individual moved during a 30-day time-bin, we created a weighted average of each environmental measure, with weights determined by the proportion of time spent at each location during the bin to select the most common address during the time. In addition to the structured features (*EHR*), 17 features were added to each time-bin as ABMH features (*EHR+ABMH*) to train the model.

### Classification Models

#### Machine Learning-based Models

For each age group, we apply ML models such as Logistic Regression (LR), Decision Tree (DT), Random Forest (RL) [28], K-Nearest Neighbor (KNN), and Extreme Gradient Boosting (XGBoost) [29]. All models are widely used in classification tasks in various domains such as image recognition [33, 34], natural language processing [35, 36], and recommender systems [37, 38]. For all ML-based models, we use the last 30-day time-bin, *x*_*T*_, from Fig 1, as an input feature to prevent the feature size from becoming too large. The increment of the feature size can cause a curse of dimensionality [39], which results in performance degradation by losing the meaning of the distances of the data points. Detailed information on each model is introduced in S2 Appendix.

#### Deep Learning-based Models

For the DL-based models, we apply Long Short-Term Memory (LSTM) [31], Gated Recurrent Unit (GRU) [30], Reverse Time Attention (RETAIN) [16], and Diagnosis Prediction Model (Dipole) [17]. Unlike ML-based models, all time-bins (*x*_1_, *x*_2_, …, *x*_*T*_ from the Fig 1) are used as input feature matrix. We use Recurrent Neural Network (RNN)-based models that are effective, especially with sequential information, to handle such a long sequence of encounters. Table 1 shows the statistics of the features and time-bins used for each age group. Note that the feature size in Table 1 is the feature size of *EHR*. Detailed information on each model is introduced in S2 Appendix.

### Experimental Setting

Model training and hyperparameter tuning are conducted to generate accurate classifications of patients at risk of developing anxiety. For hyperparameter tuning, we split the dataset into 64% training, 16% validation, and 20% test sets. All models use the same setting and are compared with the same split across the age group. All ML models are implemented using the scikit-learn package [40], except XGBoost, which has its dedicated library [29]. The implementation of LSTM and GRU uses the PyTorch library [41]. We use the source code provided by the papers for RETAIN^1^ and Dipole^2^. We optimize the ML-based models through a grid search for the most effective hyperparameters using the scikit-learn package [40]. For the DL-based models, we perform fine-tuning using the Adam optimizer with a grid search. S3 Appendix summarizes the hyperparameters used for tuning and shows the selected hyperparameters for each model. Hyperparameters are selected using the *EHR* features, and the same hyperparameter settings are used throughout the experiment.

### Evaluation Metrics

We present six metrics for the pediatric anxiety classification task in this study: (1) accuracy, (2) area under the receiver operating characteristics curve (AUROC), (3) area under the precision and recall curve (AUPRC), (4) positive predictive values (PPV), (5) negative predictive values (NPV), and (6) F1 score. The AUROC is a common evaluation tool that quantifies overall classification performance. The AUROC scale is from 0 to 1, with a value of 0.5 indicating an uninformative classifier. Higher AUROC scores typically signify better performance. While AUROC measures the area under the true positive and false positive rate, AUPRC evaluates the performance on the precision and recall graph. AUROC is a metric independent of class imbalance, and AUPRC is a metric for imbalanced datasets. The F1 score is the harmonic mean of the precision and recall of a classification model. It ranges between 0 and 1, and models with values closer to 1 are better models. PPV and NPV represent the accuracy of a diagnostic test in identifying true positive and true negative results, respectively. Predictive values indicate the likelihood that a specific diagnosis given by a test is accurate for a subject. The following equation computes PPV:

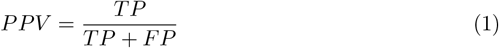

where TP is the true positive (the number of cases correctly identified as anxiety), and FP is the false positive (the number of cases incorrectly identified as anxiety). NPV can be computed with:

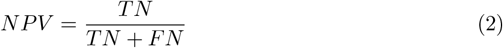

TN is the true negative (the number of cases correctly identified as non-anxiety), and FN is the false negative (the number of cases incorrectly identified as non-anxiety).

## Empirical Results

We compare the performance of ML- and DL-based models using two datasets: (1) structured features generated from EHR data (*EHR*) and (2) the combination of structured EHR and ABMH data (*EHR+ABMH*). We utilize only the final time-bin which includes the 30-days prior to the 30-day blackout period to the time of diagnosis for the ML models, whereas for the DL models, we incorporate all of the time-bins. Our model performance was bolstered by statistical analysis through 1000 bootstrapping iterations, ensuring a robust assessment of each model’s predictive capabilities. Fig 2 and Fig 3 display the mean of 1000 bootstrapping iterations at each data point, while the colored region represents the 95% confidence interval (CI). However, CIs are tight; therefore, they are not visible in some places on the figure. This section exclusively focuses on the evaluation measures AUROC score and PPV. S4 Appendix provides results of additional evaluation metrics, such as accuracy, AUPRC, NPV, and F1 score of all ML and DL models with two datasets across different age groups.

**Fig 2.**
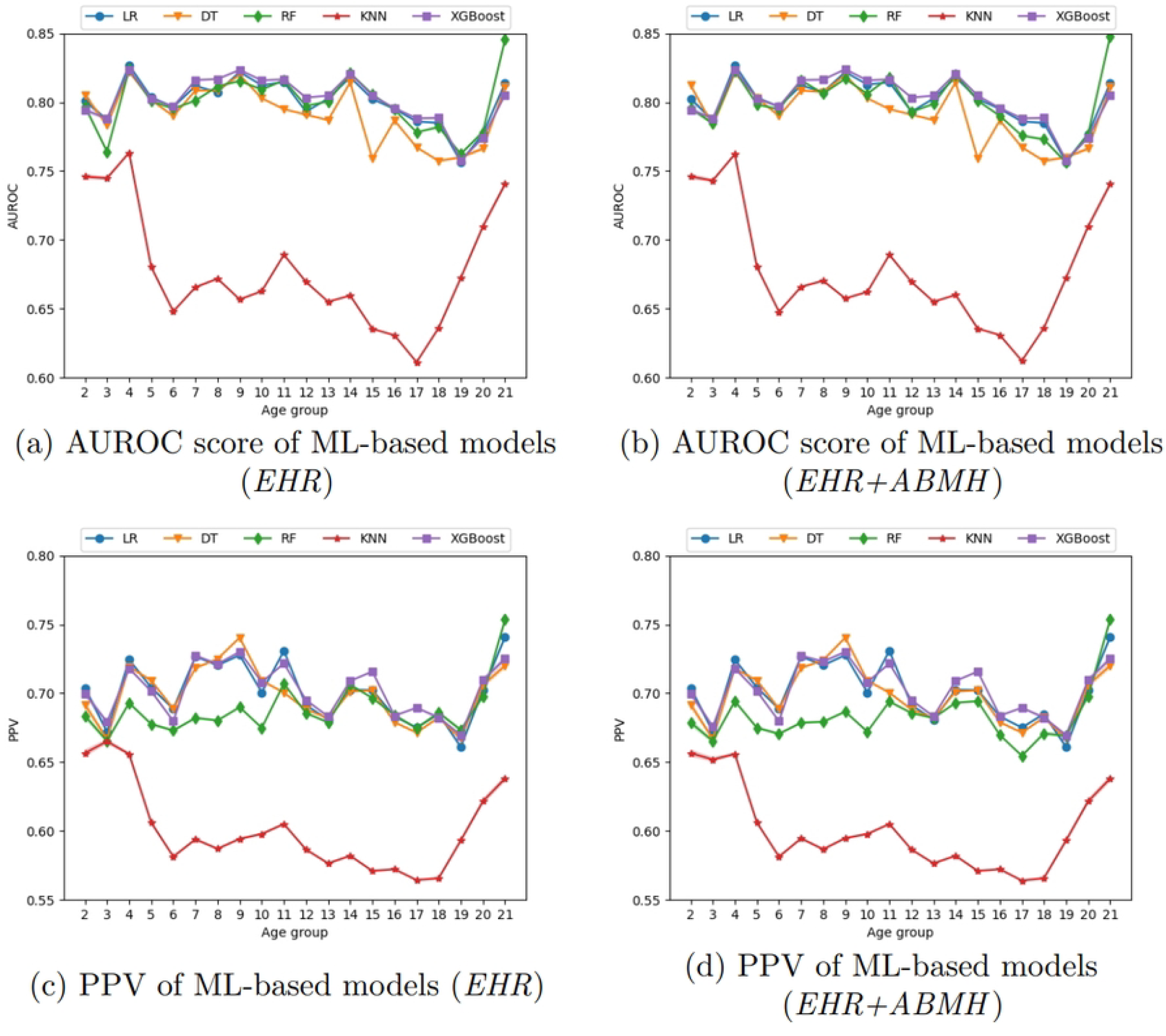
AUROC score and PPV results of ML-based models.

**Fig 3.**
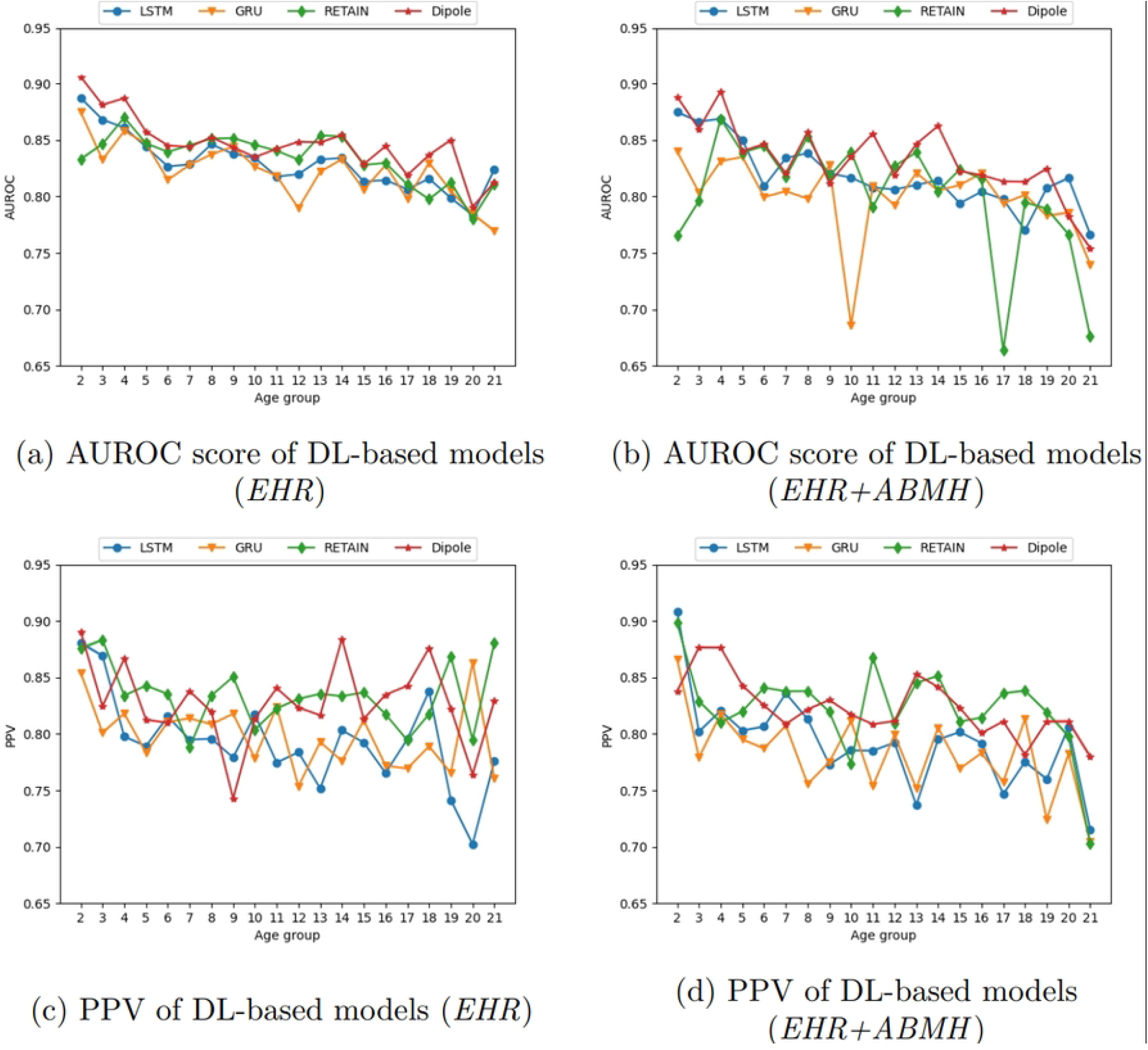
AUROC score and PPV results of DL-based models.

### Machine Learning-based Models

Fig 2 displays the outcomes of ML models in terms of Area Under the Receiver Operating Characteristic (AUROC) score and Positive Predictive Value (PPV) for all age groups. Fig 2(a) and (c) employ the *EHR* features, while Fig 2(b) and (d) showcase outcomes utilizing *EHR+ABMH* features. Fig 2(a) and (b) display the AUROC score, whereas Fig 2(c) and (d) display the PPV for each model across the age groups.

According to the results shown in Fig 2(a) and (b), XGBoost consistently beats other ML models in terms of AUROC score in most of the age groups. RF demonstrates the second-best performance, while KNN performs worse than all other models. RF surpasses XGBoost in AUROC score in age groups 2 and 21, with the lowest number of patients. Nevertheless, Fig 2(c) and (d) demonstrate that LR and DT exhibit superior PPV compared to RF. This indicates that LR and DT have a higher precision but a lower predictive performance than the other two models. The findings generally indicate that XGBoost performs better than other models in terms of AUROC score and PPV, while KNN performs worse than other models.

### Deep Learning-based Models

Fig 3 displays the outcomes of DL models in terms of AUROC score and PPV for all age groups. Fig 3(a) and >(c) employ the *EHR* features, while Fig 3(b) and >(d) display the outcomes utilizing *EHR+ABMH* features in AUROC score and PPV, respectively.

Fig 3(a) demonstrates that either RETAIN or Dipole achieve higher AUROC scores than other DL models in most of the age groups. However, in the case of the results utilizing *EHR+ABMH* features, unlike the ML results, which exhibit identical outcomes to those with *EHR* features, the results utilizing *EHR+ABMH* features display a distinct pattern. Although most models show a consistent pattern when using *EHR* features, *EHR+ABMH* features experience declines in certain age groups (namely, the GRU results for age group 10 and the RETAIN results for age group 17). Furthermore, although RETAIN demonstrates high performance in AUROC score when utilizing *EHR* features, *EHR+ABMH* features do not exhibit the same outcome. Fig 3(b) demonstrates that LSTM yields more consistent results but does not surpass other models. In both datasets, the RETAIN and Dipole models have superior AUROC scores, surpassing all other models in most age groups.

The PPV results exhibit greater complexity, as depicted in Fig 3(c) and >(d). According to the data, Dipole does not demonstrate the highest PPV across the age groups. On the contrary, RETAIN demonstrates superior PPV in about 50% of the age groups, although the AUROC score is lower than Dipole in most age groups. From this, we infer that the findings of RETAIN exhibit higher precision but a poorer predictive performance than Dipole. For some age groups, specifically 2, 3, 20, and 21, the 95% confidence interval (CI) is substantial. This means the data exhibits significant variability, which could impact the analysis. It also shows that the patient population needs to be increased to train the model adequately. For instance, for the results utilizing *EHR* features (Fig 3(c)), age group 2 consists of 930 patients, resulting in a CI of 0.0026 for the PPV of the RETAIN model. On the other hand, age group 16 includes 11,150 patients and a CI of 0.00088. This indicates that the size of the dataset is also essential to train a more effective model.

## Discussion

Our study demonstrates the considerable potential of ML- and DL-based models to classify anxiety disorders in pediatric patients using structured features generated from EHR data and the combination of structured EHR and Area-based measures of health (ABMH) data. The use of ML models can leverage the predictive ability of structured features, such as diagnosis codes, demographics, and risk assessments, to classify pediatric mental health [11, 42, 43]. Many ML classification models, such as decision trees [44, 45], support vector machines [46, 47], and random forests [48, 49] have previously been successfully used to classify anxiety disorders using structured features.

Unlike typical machine learning or ‘shallow’ learning approaches, deep learning employs artificial neural networks inspired by the structure and operation of the brain. Especially for time-dependent features, recurrent neural networks (RNNs) are introduced to handle sequential information to offer insights into disease development over time [50]. Many variants of RNN models have been proposed, for example, Long Short-Term Memory (LSTM) networks, and these models have proven highly effective in assessing time-series data, including patient visit timelines, symptom development, or treatment history [9, 10, 16, 17, 51]. These models can accurately represent the complex temporal patterns involved in the emergence and progression of anxiety disorders in pediatric patients. The ability to predict future symptoms based on previous data enables timely intervention, leading to substantial improvements in patient outcomes.

ABMH data provides information about a child’s social and physical environment, which is incorporated into ML and DL models [52–54]. ABMH data contain residential history and environmental information to help understand pediatric anxiety. Incorporating these determinants into ML and DL models can improve the prediction power of the anxiety classification models and address health disparities [55, 56]. However, when constructing the ABMH data, we assumed that the patients resided at the same location between clinic visits and that any address changes were effective starting at the date of the visit. There is potential for bias in this assumption, as patients may move locations between visits, which may artificially affect the impact of the ABMH measures in our models.

Anxiety disorders develop differently in different age groups and also depend on the social and physical environment of each individual. For example, preschool-aged children may develop anxiety disorders when they are separated from their primary caregivers or experience traumatic separation-related events [57], school-aged children may develop anxiety disorders due to their certain school environment [58], and adolescents may develop anxiety disorders through certain psychological processes which may be perpetuated or modeled in certain family environment [23]. Understanding these age-specific manifestations is essential for accurate diagnoses and appropriate treatment.

## Conclusion

The findings of this study affirm that employing ML and DL models can enable the identification of age-stratified pediatric patients at high risk of anxiety onset, although parametric logistic regression (LR) models performed at least as well as other ML-based models (DT, RF, KNN, XGBoost). The consistency of the models and predictive strength have substantial implications for enhancing clinical decision-making and patient outcomes. Integrating such models into healthcare practices promises a shift toward more efficient, data-driven, and personalized care. Future efforts should focus on customizing these models for diverse patient cohorts to maximize their utility in real-world settings and compare the age-stratified model to an all-age one. This study underscores the potential of a data-driven methodology, streamlining early detection and catalyzing a transformative shift in pediatric mental healthcare practices.

## Data Availability

Data cannot be shared publicly because it contains personally identifiable information and would be protected health information as such, we are not authorized to make our dataset publicly available. This data was acquired and used under an approved IRB protocol from Cincinnati Children's Hospital Medical Center. Data are available from the Cincinnati Children's Hospital Medical Center Institutional Data Access / Ethics Committee for researchers who meet the criteria for access to confidential data.

## Supporting information

**S1 Table. List of ICD codes to determine anxiety patients**.

**S2 Table. Area-based Measures of Health (ABMH) data description**.

**S1 Appendix. Structured EHR Data**. Provide extensive information about the time-dependent and static features in the dataset.

**S2 Appendix. Classification Models**. Discuss the specifics of the ML- and DL-based classification models that are applied.

**S3 Appendix. Hyperparameter Tuning**. Discuss the hyperparameter tuning procedure, as well as the hyperparameters selected for ML- and DL-based models.

**S4 Appendix. Additional Results**. Present the additional results using various evaluation metrics (accuracy, NPV, F1 score, and AUPRC) for both *EHR* and *EHR+ABMH* features of ML- and DL-based models.

## Acknowledgments

This work was supported by Cincinnati Children’s Hospital Medical Center under Strategic Partnership Projects agreement NFE-21-08617. This work has been authored by UT-Battelle, LLC under Contract No. DE-AC05-00OR22725 with the U.S. Department of Energy. This research used resources of the Oak Ridge Leadership Computing Facility, which is a DOE Office of Science User Facility supported under Contract DEAC05-00OR22725.

## Footnotes

1 https://pyhealth.readthedocs.io/en/latest/_modules/pyhealth/models/retain.html#RETAINLayer

2 https://github.com/Yuyoo/Dipole/blob/master/dipole_torch/Dipole_torch.py

